# ACE2 Netlas: In-silico functional characterization and drug-gene interactions of *ACE2* gene network to understand its potential involvement in COVID-19 susceptibility

**DOI:** 10.1101/2020.10.27.20220665

**Authors:** Gita A Pathak, Frank R Wendt, Aranyak Goswami, Flavio De Angelis, COVID-19 Human Genetics Initiative, Renato Polimanti

## Abstract

Angiotensin-converting enzyme-2 (*ACE2*) receptor has been identified as the key adhesion molecule for the transmission of the SARS-CoV-2. However, there is no evidence that human genetic variation in ACE2 is singularly responsible for COVID-19 susceptibility. Therefore, we performed a multi-level characterization of genes that interact with ACE2 (ACE2-gene network) for their over-represented biological properties in the context of COVID-19.

The phenome-wide association of 51 genes including ACE2 with 4,756 traits categorized into 26 phenotype categories, showed enrichment of immunological, respiratory, environmental, skeletal, dermatological, and metabolic domains (p<4e-4). Transcriptomic regulation of ACE2-gene network was enriched for tissue-specificity in kidney, small intestine, and colon (p<4.7e-4). Leveraging the drug-gene interaction database we identified 47 drugs, including dexamethasone and spironolactone, among others.

Considering genetic variants within ± 10 kb of ACE2-network genes we characterized functional consequences (among others) using miRNA binding-site targets. MiRNAs affected by ACE2-network variants revealed statistical over-representation of inflammation, aging, diabetes, and heart conditions. With respect to variants mapped to the ACE2-network, we observed COVID-19 related associations in *RORA, SLC12A6* and *SLC6A19* genes.

Overall, functional characterization of ACE2-gene network highlights several potential mechanisms in COVID-19 susceptibility. The data can also be accessed at https://gpwhiz.github.io/ACE2Netlas/

## 1 Introduction

SARS-CoV-2 (severe acute respiratory syndrome coronavirus 2) is the causative agent responsible for recent global spread of COVID-19 (coronavirus disease 2019) [1,2]. Millions of people have been infected with the virus, which caused global lockdowns and heavily restricted interpersonal contact. These measures were taken to reduce viral spread through respiratory droplet exchange between persons.

SARS-CoV-2 is capable of entering the host cells via ACE2 (angiotensin converting enzyme 2) [3]. ACE2 is found on many different cell types, which normally helps regulate blood pressure and inflammation through cleavage of angiotensin II (ANG II) [4]. The virus occupies cell-surface of *ACE2* leading to accumulation of angiotensin (ANGII), inflammation, and cell death [3]. In the lungs, SARS-CoV-2 mediated ANGII accumulation leads to alveolar cell death and a reduction in oxygen uptake [5].

Although ACE2 is the cellular entry point, there is little evidence that genetic variation in *ACE2* is singularly responsible for COVID-19 susceptibility. Indeed, *ACE2* failed to associate with COVID-19 informative phenotype definitions from large genome-wide studies [6–8]. However, due to the functional role of ACE2 in SARS-CoV-2 infection, we hypothesize that genes interacting with ACE2 activity are enriched for molecular pathways relevant for COVID-19 susceptibility. Accordingly, we employed a top-down approach to analyze tissue-specific transcriptomic regulation, drug-gene interactions, and variant prioritization using genetic variants within the ACE2 gene-gene connectome and protein-protein interaction networks. With this approach we identified several biological processes and functional effects of ACE2-gene network relevant for the vast symptoms observed following SARS-CoV-2 infection.

## 2 Results

A study overview is presented in Supplementary file1 Figure S1.

### 2.1 The ACE2 gene connectome

A total of 60 ACE2-interacting genes were identified from different network databases (Supplementary file2 Table S1; Figure 1).

**Figure 1:**
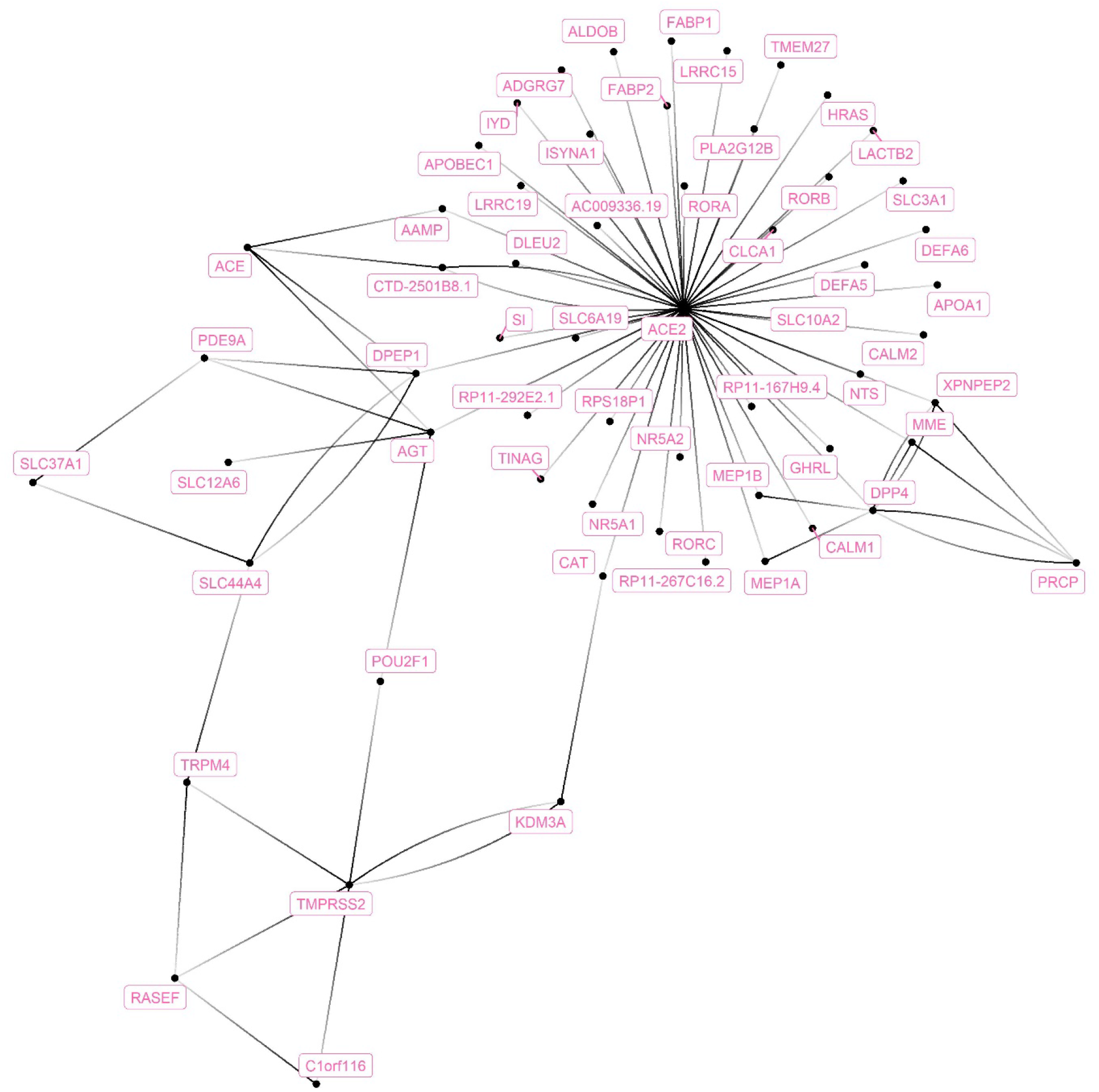
The ACE2-gene network. The genes that connect with ACE2 were extracted from six different gene-network databases and compiled together in one network.

### 2.2 Tissue-specific transcriptomic regulation

Using differential expression data of 54 tissues (GTEx-v8), the genes in the ACE2-gene network were enriched for upregulated expression in small intestine (p=1.07×10^−16^), colon (p =7.60×10^−13^), kidney (p=1.93×10^−8^), and liver (p=4.63×10^−4^) (Figure 2; Supplementary file2 Table S2). No tissue-specific enrichment was observed for down-regulated expression.

**Figure 2:**
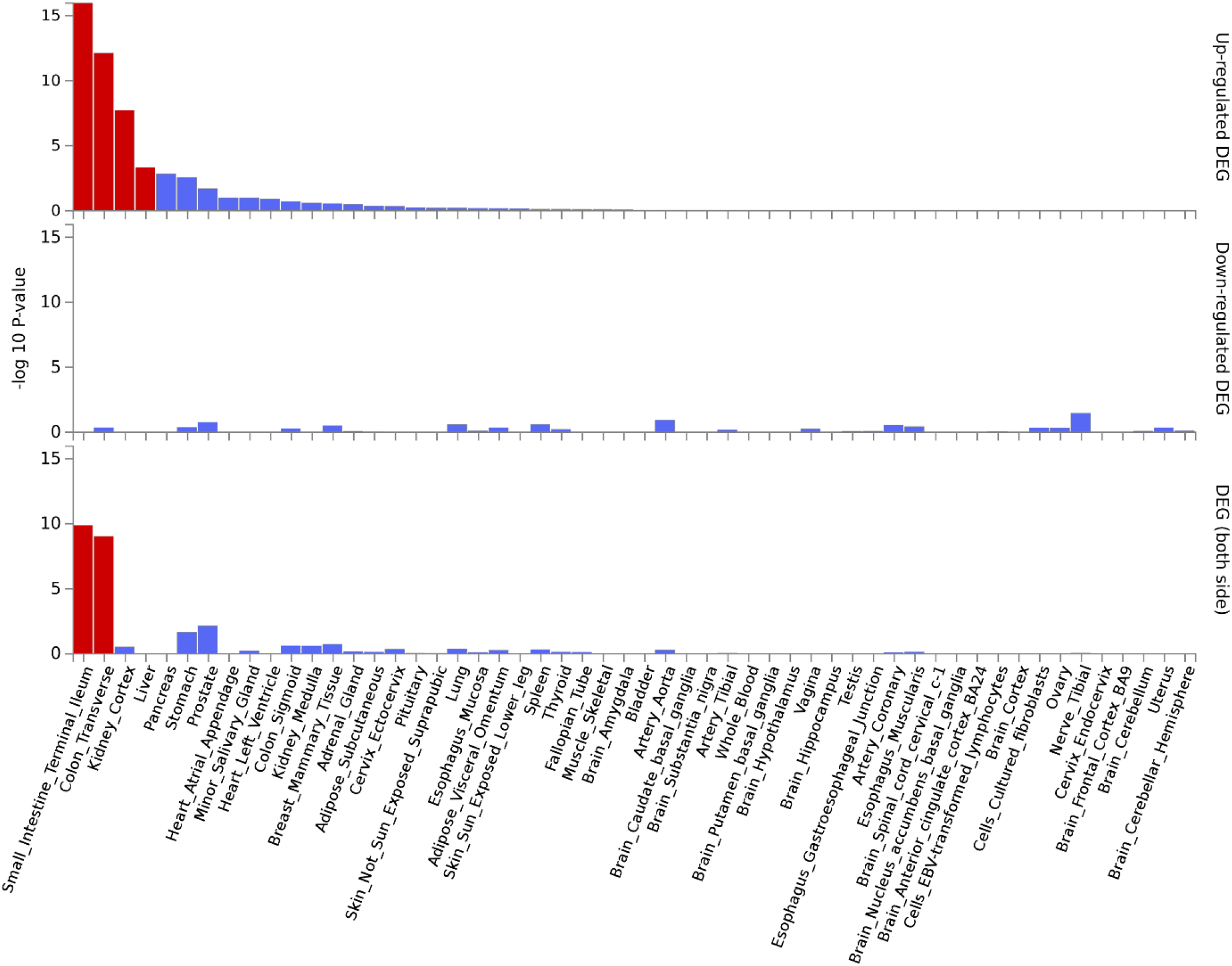
Tissues enriched based on ACE2-network gene expression for GTEx tissues. The genes from the ACE2-network show over-representation of tissues (x-axis) and -log10 p-value (y-axis). The red bars are significant enrichments.

### 2.3 Gene-Drug Interaction and Over-represented Biological Functions

Out of 61 genes, 29 had information about their drug-gene interaction in the drug-gene interaction database (DGIdb)[9]. This assessment resulted in 238 unique drug-gene observations (Supplementary file2 Table S3). Some of the notable drugs observed via this approach were spironolactone, dexamethasone, metformin, and hydrocortisone. To understand the role of these drugs in affecting biological processes, we performed drug-set enrichment analysis. DSEA [10] found gene-ontology mapping for 47 drugs and tested against REACTOME gene ontology database. Although the results did not survive Bonferroni correction, the strongest enrichments were observed for platelet sensitization by low-density lipoprotein cholesterol (p=0.003), IL-7 signaling (p=0.004), glycerophospholipid biosynthesis (p=0.005), and viral messenger RNA synthesis (p=0.011) (Figure 3; Supplementary file2 Table S4).

**Figure 3:**
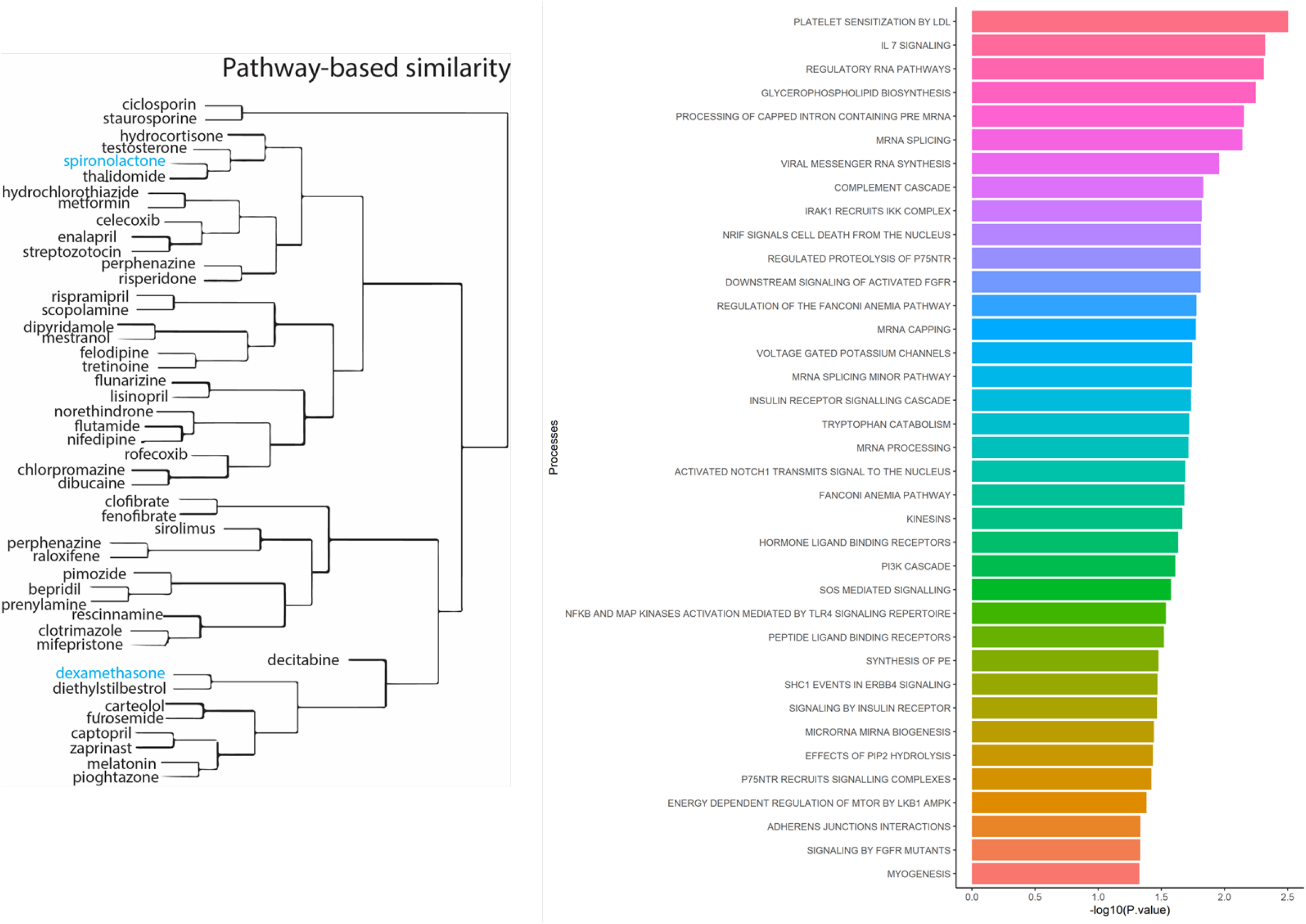
Drug-set enrichment analysis. LEFT: The similarity of drugs based on pathways identified. RIGHT: Biological Processes identified based on drugs that interact with genes from the ACE2-network.

### 2.4 Over-representation of phenotypic domains within ACE2 gene network

A phenome-wide association study (PheWAS) was performed for 51 genes leveraging data from the GWASAtlas [11]. The GWASAtlas categorized traits into 26 phenotype domains (Supplementary file1 Figures S2-S52; Supplementary file2 Table S5). Each domain was tested for enrichment of significant traits versus non-significant traits (Supplementary file2 Table S6). Six domains were significant: ‘Immunological’ (p=7.62×10^−25^), ‘Respiratory’ (p=1.30×10^−8^), ‘Skeletal’ (2.94×10^−8^), ‘Dermatological’ (p=7.91×10^−8^), ‘Environmental’ (p=2.21×10^−7^), and ‘Metabolic’ (4.33×10^−4^) (Supplementary file2 Table S7). *SLC44A4* had the highest number of associated traits across the significant domains (n_total_= 173) followed by *APOA1* had highest number of traits associations, mostly metabolic (n_total_= 100; metabolic = 71) (Figure 4). *SLC44A4, APOA1*, and *RORA* showed associations across all six enriched domains.

**Figure 4:**
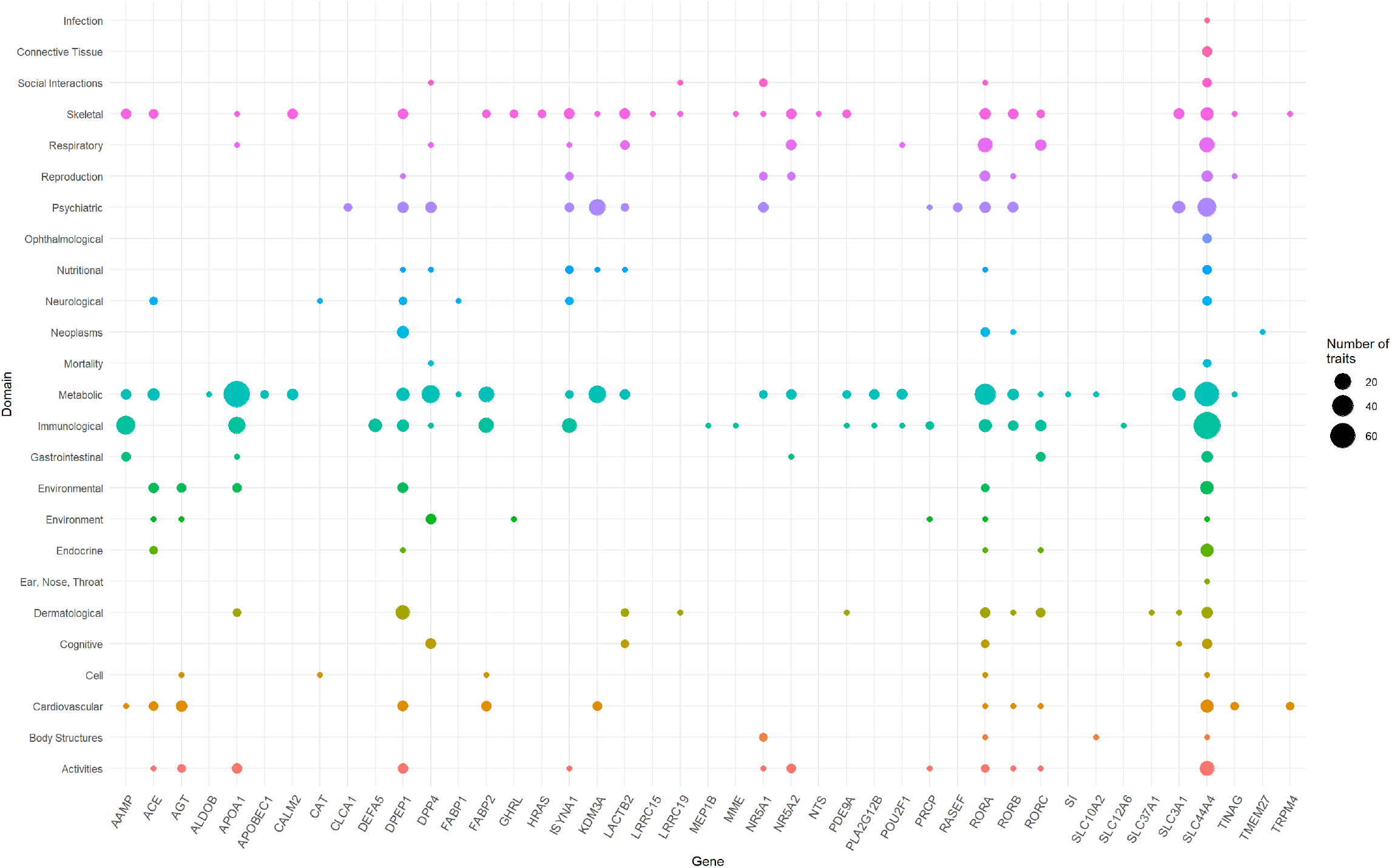
Domain distribution of PheWAS of ACE2-network genes. The ACE2 gene network associations are grouped based on domains (y-axis) and gene names (x-axis). The size of the data points reflects number of phenotypes surviving multiple testing correction.

### 2.5 Characterization of SNPs

We extracted all 957,222 SNPs in the ACE2-network and annotated for allele frequency (Supplementary file3), nearby genes and coordinates (Supplementary file4), Combined Annotation Dependent Depletion (CADD) [12] and DeepSEA [13] scores. There were 98,529 SNPs with CADD score >10, which corresponds to the top 10% pathogenic variants across the human genome (Supplementary file5). To identify their regulatory consequences, variants were annotated with DeepSEA which provides functional probability of the SNPs in serving as gene expression, disease and chromatin regulating variants. There were 12,095 SNPs within the ACE2-gene network which had >50% functional probability (DeepSEA functional score > 0.5) (Supplementary file6). The miRNAs altered by the SNPs were analyzed for over-represented miRNA-family, biological functions, and diseases considering false discovery rate multiple testing correction (FDR p<0.05). There were 4 miRNA clusters that were enriched, miR-302b, miR-181d (p=0.0079), miR-17, and 106a (p= 0.00298). We found 65 biological functions that were significant and the top five significant biological processes were cell death (p=1.5×10^−20^), inflammation (p=2.57×10^−20^), cell cycle (p=2.09×10^−18^), apoptosis (p=4.15×10^−18^), and immune response (p=3.17×10^−17^) (Figure 5). We observed a total of 152 significant diseases of which the most significant were diabetes mellitus type 2 (p=1.55×10^−22^), hepatitis c virus infection (p=5.56×10^−21^), atherosclerosis (p=3.08×10^−19^), heart failure (p=4.22×10^−19^), and Alzheimer’ s disease (p=1.35×10^−17^) (Supplementary file2 Table S8).

**Figure 5:**
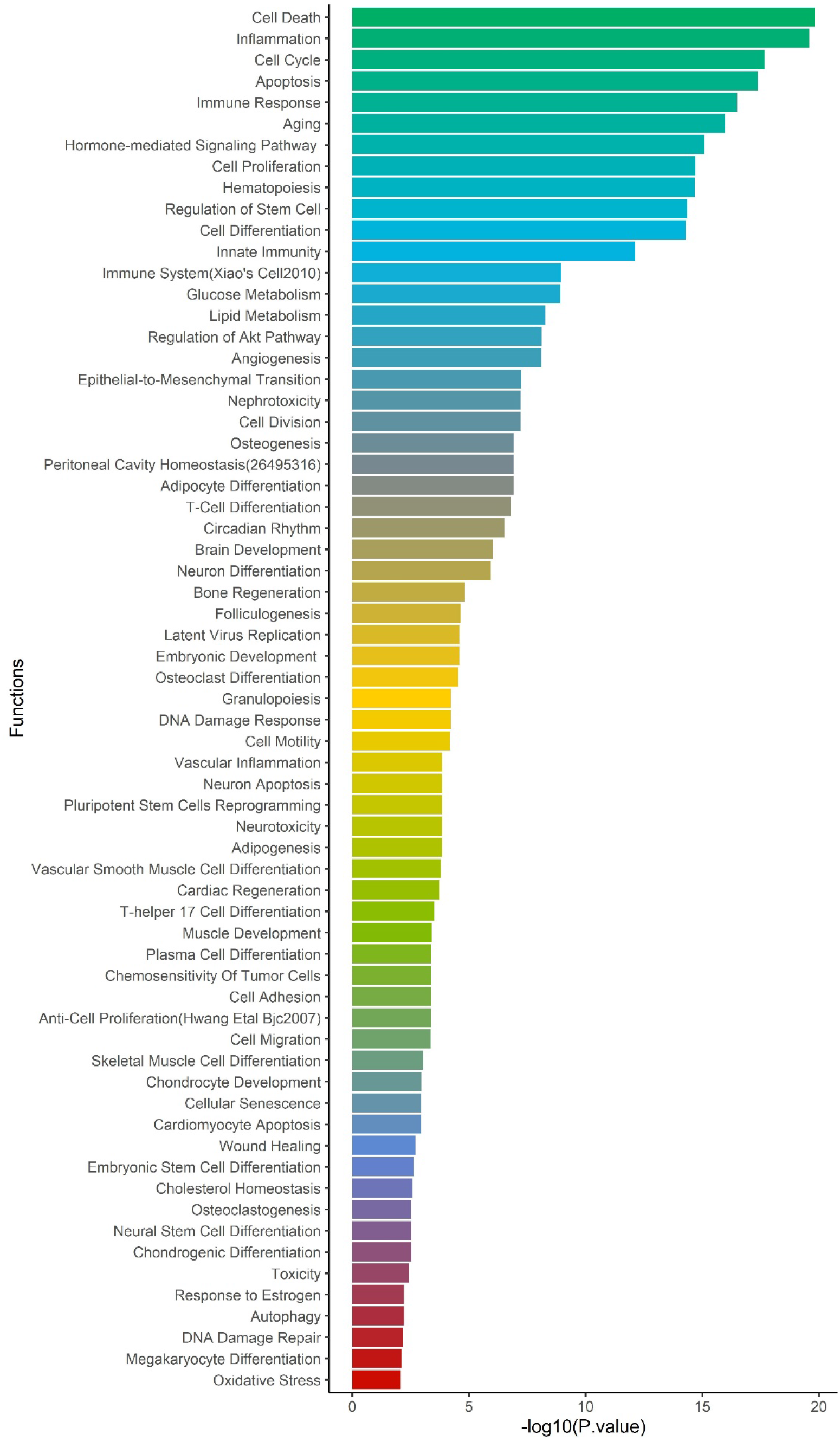
Enrichment of biological functions based on miRNA:SNP annotation. Using miRNAs annotation, over-represented biological processes are shown on y-axis and -log10 pvalue on x-axis.

### 2.6 Neanderthal LA introgression within ACE2 network SNPs

Due to the Neanderthal introgression observed in 3p21 locus as risk to COVID-19 [14], we compared mean probability of Neanderthal LA between the ACE2-network SNP set (mean=0.032) and 1,000 randomly selected SNP sets with comparable genomic features (range of Neanderthal LA means = 0.027-0.036). The ACE2-network SNPs did not show evidence of Neanderthal LA introgression significantly different from those expected by chance (p=0.663) (Supplementary file1 Figure 55).

### 2.7 Annotation of network SNPs using the COVID-19 GWAS

We tested ACE2-network SNPs with respect to six COVID-19-related phenotypes (Freeze 3) released by the COVID-19 Host Genetics Initiative [15]. To identify independent variants, the variants were pruned for linkage disequilibrium (LD<0.1 within 250kb genomic size) and clumped for p-value <0.01. Variants surviving multiple testing were annotated for eQTLs, and mQTLs. Three genes – *RORA, SLC12A6*, and *SLC6A19* – showed associations with multiple COVID-19 phenotypes (Supplementary file2 Tables S9-S14; Supplementary file6 Figures S56-S61). *RORA* SNPs were associated with COVID-19 positive status (rs17303202, p=2.35E-5), laboratory-confirmed positive COVID-19 status (rs4774377, p=8.25E-5), hospitalized COVID-19 (rs17303202, p=2.76E-05), and COVID-19 with very severe respiratory symptoms (rs341419, p=8.13E-4). The SNPs in *RORA* gene are also associated with gene expression of *RORA* gene (rs12912196; p=3.9E-5) and trans-mQTL (cg00930615 in *ANXA2*). *SLC12A6* associations were observed with respect to COVID-19 (rs145719616, p=1.19E-4), hospitalized COVID-19 (rs192235418, p=4.42E-4), COVID-19 with very severe respiratory (rs2705343, p=1.86E-3), and. *SLC6A19* SNPs were associated with severe COVID-19 phenotype definitions, i.e. COVID-19 with very severe respiratory confirmed (rs76067074, p=2.65E-3) and hospitalized COVID-19(rs76067074, p=2.52E-4).

## 3 Discussion

*ACE2* is expressed in several tissues and plays a key role in host-entry of SARS-CoV-2 [16]. However, the genomic profile of *ACE2* is limited in explaining the vast symptomology observed for COVID-19. Understanding ACE2 associated molecular networks presents several functional insights between genetic targets based on gene expression, topology, and protein and signaling relationships [17]. Due to the well-characterized role of ACE2 in SARS-CoV-2 infection, we generated novel information regarding the molecular and phenotypic characteristics of ACE gene network in the context of their potential involvement in COVID-19 susceptibility. Our PheWAS-based analysis showed that genetic variation within ACE2 gene network is associated with immunity, respiratory, and metabolic traits. This is in line with known epidemiology of COVID-19 and its comorbidities [18,19].

The expression of ACE2-network genes was enriched for regulatory mechanisms related to small intestine, colon, kidney, and liver. It is hypothesized that furin, a serine protease present in lungs but also highly expressed in small intestine, presents S-spike for attachment of the ACE2 receptor [20]. Patients with kidney disease have higher risk for COVID-19 severe symptoms [21]. Additionally, the inflammation and cytokine storm from COVID-19 is observed to damage kidney tissues [22]. Lastly, modest increase in liver enzymes has been associated with COVID-19, and returning to baseline during the recovery phase [23].

Understanding the genes that interact with *ACE2* receptor has potential to understand drug-targets and molecular processes that might play a role in susceptibility and treatment response of COVID-19. The drug-gene interaction analysis within ACE2 network identified dexamethasone, reported to lower mortality in COVID-19 cases requiring mechanical ventilation [24]. Drugs — spironolactone and hydrocortisone target the androgen system. The androgen receptor has been associated with severe symptomology of COVID-19 [25]. Spironolactone is a diuretic and alleviates respiratory symptoms by reducing fluid from the lungs [26]. The use of spironolactone is currently being tested for acute respiratory distress syndrome in COVID-19 patients [27]. Hydrocortisone is currently under clinical trials for treating COVID-19 related hypoxia symptoms [28]. Among the other compounds identified, metformin, a known drug for treating diabetes, can also affect respiratory outcomes [29]. A recent study reported protective effects of metformin in women with diabetes and obesity who were admitted with COVID-19 diagnosis [30]. Lastly, melatonin has been hypothesized to improve general immunity and lower oxidative stress generated from SARS-CoV-2 infection [31].

The miRNA target sites altered by ACE2-network SNPs identified miR-302b and miR-181d as over-represented miRNA clusters. The downregulated expression of miR-302b has been observed to reduce survival rates in chronic obstructive pulmonary disease (COPD) patients [32]. A meta-analysis showed that COPD diagnosis increased susceptibility to COVID-19 [33]. The miRNA-181 cluster has been associated with regulation of TNF-alpha [34], T-cell aging [35] and emphysema [36]. miRNA-17 and 106 belong to same miRNA family, miRNA-17 is upregulated in bronchoalveolar stem cells to lower SARS-CoV replication [37]. An *in silico* study of miRNA targets for SARS-CoV-2 genomic sequence found miRNA-17 as one of the targets with experimental evidence of its upregulation in H7N9 Influenza virus infection [38]. The top over-represented diseases in miRNA-ACE2-network-SNPs were diabetes, hepatitis C viral infection, heart failure and Alzheimer’ s disease. COVID-19 in individuals with diabetes has been reported to require hospitalization than non-diabetic individuals [39]. Furthermore, SARS-CoV-2 infection contributes in the development of ketosis in diabetic individuals resulting in longer length of hospitalization stay [40]. Triglyceride and glucose index was associated with severity of COVID-19 [41]. While there are limited studies about hepatitis C in COVID-19 patients [42], heart failure was reported by multiple studies as being associated with COVID-19 severity [43,44]. Alzheimer’ s disease is another condition associated with COVID-19 susceptibility [45], including *APOE4* carrier status with increased risk of severe COVID-19 [46].

In contrast to specific enrichment of Neanderthal LA in a COVID-19 risk locus on chromosome 3 [47], there is no evidence of increased Neanderthal LA in the ACE2 network investigated here. This suggests that, although some loci conferring risk for COVID-19 severity, such as the one identified on chromosome 3, may have originated from Neanderthal admixture events, this mechanism did not shape the genetic architecture of the ACE2 network responsible for entry of SARS-CoV-2 into host cellular machinery.

Lastly, among ACE2-network-SNPs, potential COVID-19 risk alleles were observed in *RORA* gene with respect to multiple COVID-19 phenotypes. *RORA* protein product is involved in immune response, cancer and metabolism [48]. *RORA* plays a role in the activation of T helper cells during lung inflammation by regulating tumor necrosis factor and interleukins [49,50], and its protein product showed multiple regulatory functions in human epithelial cell cultures inoculated with SARS-CoV-1 [51]. The hypothesis-free approach of genome-wide association of hospitalized COVID-19 vs the population highlighted *SLC6A20* with genome-wide significance on chromosome 3 locus. The SLC12 (*SLC12A6*) class is responsible for inorganic ions such as sodium and chloride while the SLC6 class (*SLC6A19*, identified via network approach and SLC6A20, identified via genome-wide approach) are responsible for transport of amino acids such as glutamate and glycine which are important neurotransmitter activity [52]. *SLC6A19* (among other SLC-class genes) serves similar function to *SLC6A20*, both are expressed in the intestinal tissue and contingent upon ACE2 expression [53]. Multiple studies report more than 10% of the COVID-19 confirmed patients exhibit gastrointestinal symptoms[54– 56]

Although we provided a wide range of information highlighting the molecular and phenotypic characteristics of *ACE2* gene network and their putative implications with COVID-19 risk, the findings reported have to be considered exploratory. We used appropriate computational methods and statistical approaches to generate reliable evidence useful to open new directions in COVID-19 research. We also highlighted when the results reported did not survive stringent multiple testing correction. This limitation is particularly relevant with respect to the ACE2 network genetic associations. Due to the limited statistical power of the genome-wide data available to date, none of the risk alleles identified as functionally relevant survive genome-wide testing correction. Further analyses will be needed to validate our current findings.

## 4 Conclusion

*ACE2* is one of the few molecular targets recognized to play a key role in the COVID-19 pathogenesis. We conducted a comprehensive analysis leveraging multiple resources (e.g., drug-gene interactions, tissue-specific transcriptomic profile, and phenome-wide and genome-wide datasets) to expand our understanding of the genomic characteristics of the host *ACE2* gene network. Overall, our findings highlight the potential mechanisms linking *ACE2* systems biology to COVID-19 susceptibility.

## 5 Methods

### 5.1 Gene network collection

Information regarding ACE2 gene network was mined from GeneMANIA [57], Stringdb [58], APID [59], GeneNetwork [60], Biogrid[61] and FunctionalNet [62]. Immediate genes connections that were available in each databank with their default settings result in 61 unique genes (60 genes plus *ACE2*) (Supplementary file1 Figure 1; Supplementary file2 Table S1). The genomic coordinates for the genes were annotated using biomart [63], ensemble GRCh37/hg19. The analysis and visualization were performed in R 3.6.

### 5.2 2.2 Tissue-specific transcriptomic regulation

The tissue specificity was tested for 60 ACE2-interacting genes in FUMA [64]. The input genes were tested for pre-calculated tissue-specific differentially expressed genes from the GTEx v8 [65]. We also considered the t-statistic sign for up and down-regulated genes against protein coding genes as background. Enrichments were performed using hypergeometric tests and significant enrichments were defined according to Bonferroni corrected p-value ≤ 0.05.

### 5.3 Phenome-wide analysis of ACE2 gene network

A phenome-wide association study (PheWAS) was performed for 51 of 60 genes that were present in GWASAtlas [11] using all traits available per gene. Statistical significance was determined by applying a Bonferroni multiple-testing correction accounting for the number of GWAS traits (4,765 traits) available in the GWASAtlas (p<1.05 x10^−5^). Each trait was grouped into a domain (Supplementary file2 Table S5) which was tested for enrichment using one-sided Fisher’ s exact test for high proportion of significant traits versus all others tested. A significant domain enrichment was defined considering a Bonferroni-corrected threshold accounting for the number of domains tested (p-value < 0.0019; 0.05/26).

### 5.4 Gene-Drug Interactions and Biological Functions

Information on drugs that interact with ACE2 network genes were extracted from The Drug-Gene Interaction database (DGIdb) [9] followed by drug-set enrichment for over represented biological functions using DSEA (Drug-Set Enrichment Analysis) [10].

### 5.5 Characterization of SNPs

Single nucleotide polymorphism (SNPs) were extracted based on the genomic coordinates of the genes (± 10kb) for GrCh37; dbSNP153 from the UCSC browser [66] using bigbed utilities [67], and referred to as ‘ACE2-network SNPs.’ ACE2-network SNPs were annotated for global allele frequency, Combined Annotation-Dependent Depletion (CADD) score [12], deep learning based algorithm framework (DeepSEA) [13], and target miRNAs using SNPnexus [68]. DeepSEA is a deep learning-based algorithmic framework for predicting the chromatin effects of sequence alterations with single nucleotide sensitivity[13].The identified miRNAs were tested for over-represented miRNA clusters, functions, and diseases using TAM 2.0 [69].

### 5.6 Neanderthal introgression

Motivated by evidence of a chromosome 3 COVID-19 risk locus enriched of Neanderthal local ancestry (LA) [47], we compared the distribution of probability of Neanderthal LA in our COVID-19 ACE2-network SNP set and 1,000 randomly sampled SNP sets comprised on SNPs across the genome with comparable genomic features. ACE2-network SNPs were mapped using previously-defined Neanderthal LA data [70,71]. A total of 6,822 LD-independent pairwise SNPs (*r*^*2*^=0.1 and *p*=0.1 in 250kb window size) were used as standard input for SNPsnap [72]. In SNPsnap, 1,249/6,822 independent ACE2 network SNPs could be matched based on the following genomic features relative to the input SNP list: minor allele frequency within 2%, gene density within 50%, nearest gene within 50%, and number of linkage disequilibrium groups within 50%. SNPsnap was instructed to exclude the ACE2-network SNP list from the pool of eligible feature-matched SNPs. Non-parametric Wilcoxon rank sum tests were used to compare the Neanderthal LA of our ACE2 network SNP list to that of all 1,000 random SNP sets and multiple testing correction was applied to adjust for a false discovery rate of 5%.

### 5.7 Association statistics of ACE2 network SNPs from the COVID-19 Host Genetics Initiative (HGI)

The ACE2-network SNPs were extracted from association statistics released by the COVID-19 HGI [15] for six phenotypes describing COVID-19 susceptibility. These phenotypes were A2_V2 (very severe respiratory confirmed COVID-19 cases [N=536] vs. population[N=329391]), B1_V2 (hospitalized COVID-19 cases [N=928] vs. not hospitalized COVID-19 cases [N=2028]), B2_V2 (hospitalized COVID-19 cases [N=3199] vs. population [N=897488]), C1_V2 (COVID-19 cases [N=3523] vs. lab/self-reported negative [N=36634]), C2_V2 (COVID-19 cases [N=6696] vs. population [N=1073072]), and D1_V2 (predicted COVID-19 cases from self-reported symptoms [N=1865] vs. predicted or self-reported non-COVID-19 cases [N=29174]). The SNPs of the ACE2 network were extracted and pruned for LD and p-value using plink1.9. The multiple testing correction was applied using Bonferroni p-value < 0.05. These significant SNPs were annotated further for pathogenicity using Combined Annotation Dependent Depletion (CADD) score and their role as quantitative trait loci (QTL) for gene expression using GTEx, and methylation using QTLbase [73].

## Data Availability

The data presented is available in above specified link.

https://gpwhiz.github.io/ACE2Netlas/

## 6 Author Contribution

G.A.P conceptualized the study design, analyzed, and drafted the manuscript. F.R.W. contributed to analysis, and manuscript writing. Authors, A.G., F.D.A. and R.P. contributed to result interpretation, manuscript drafting and revision. R.P. supervised the study and finalized the manuscript.

## 8 Acknowledgements

We would like to acknowledge support from the National Institutes of Health (R21 DC018098, R21 DA047527, R01 DA12690, F32 MH122058), and thank the COVID-19 Host Genetics Initiative (https://www.covid19hg.org/acknowledgements/) for providing open access to genetic association data.

## 9 Data Availability

The data presented is available in supplementary files and also on https://gpwhiz.github.io/ACE2Netlas/

## Figures and Supplementary file Legends

### Supplementary files

<u>Supplementary file1:</u> File containing figures S1-S60

<u>Supplementary file2:</u> Tables S1:S14. Tabular details of the list of genes, tissue enrichment, gene-drug interaction, drug-set enrichment, PheWAS of all genes, Significant traits of PheWAS, domain enrichment, miRNA enrichment, SNPs from the network for six COVID-19 phenotypes from COVID-19 Host Genetics Initiative – Freeze3

<u>Supplementary file3:</u> Text file of all the SNPs within ±10kb of the genes and their genomic coordinates and allele frequency.

- Column Headers:
  - Variation ID: <dbsnp rs#>
  - dbSNP: link to dbSNP, if known
  - Chromosome: Variant mapped chromosome location
  - Position: Variant start position on chromosome
  - REF Allele: Reference allele
  - ALT Allele (IUPAC): Observed allele
  - Minor Allele: Minor allele observed in global population, if known
  - Minor Allele Frequency: Minor allele frequency observed in global population, if known
  - Contig: Variant mapped contig location
  - contigPosition: Variant start position on contig
  - Band: SNP cytogenetic location

<u>Supplementary file4:</u> Text file of SNPs with their overlapping and nearest gene annotation using Ensembl GRCh37.

- Column Headers:
  - Variation ID: <dbsnp rs#>
  - Chromosome: Variant mapped chromosome location
  - Position: Variant start position on chromosome
  - Overlapped Gene: Name of the gene (HGNC system) to which the variant is overlapped
  - Type: Gene type, e.g., protein coding, miRNA, non coding, Pseudogene, snoRNA, lincRNA etc.
  - Annotation: Summary of whether the variant overlapped with the coding, intronic or untranslated regions of the various transcript isoforms of the gene, as annotated from Ensembl gene system.
  - Nearest Upstream Gene: If variant is not overlapped with any gene, then the gene whose end position is nearest to the variant on the left (considering the alignment of genes on the positive strand as left-to-right)
  - Type of Nearest Upstream Gene: Gene type, e.g., protein coding, miRNA, non coding, Pseudogene, snoRNA, lincRNA etc.
  - Distance to Nearest Upstream Gene: distance from the end position of the nearest upstream gene.
  - Nearest Downstream Gene: If variant is not overlapped with any gene, then the gene whose start position is nearest to the variant on the right (considering the alignment of genes on the positive strand as left-to-right)
  - Type of Nearest Downstream Gene: Gene type, e.g., protein coding, miRNA, non coding, Pseudogene, snoRNA, lincRNA etc.
  - Distance to Nearest Downstream Gene: distance from the start position of the nearest downstream gene.

<u>Supplementary file5:</u> Text file of all the SNPs with CADD (Combined Annotation Dependent Depletion) scores >10

- Column Headers:
  - Variation ID: <dbsnp rs#>
  - Chromosome: Chromosome name
  - Position: Variant start position on chromosome
  - Variant: <reference allele,”/”,observed allele> as reported in the tool’ s genome-wide score
  - PHRED: PHRED-like (−10*log10(rank/total)) scaled CADD-score ranking a variant relative to all possible substitutions of the human genome. A score≥10 indicates that it is predicted to be in the 10% most deleterious substitutions that you can do to the human genome, a score≥20 indicates the 1% most deleterious and so on.

<u>Supplementary file6:</u> Text file of all the SNPs with DeepSEA (deep learning based algorithm framework) functional scores > 0.5, which represents atleast 50% probability to have regulatory effect

- Column Headers:
  - Variation ID: <dbsnp rs#>
  - Chromosome: Chromosome name
  - Position: Variant start position in the chromosome
  - Variant: <reference allele,”/”,observed allele> as reported in the tool’ s genome-wide score
  - eQTL Probability: The probability of the variant being a eQTL variant given by functional variant prioritization classifier.
  - GWAS Probability: The probability of the variant being a trait-associated (GWAS) variant given by functional variant prioritization classifier.
  - HGMD Probability: The probability of the variant being a inherited disease-associated (HGMD) variant given by functional variant prioritization classifier.
  - Functional Significance Score: A measure in the range [0-1] depicting the significance of magnitude of predicted chromatin effect and evolutionary conservation. Lower score indicates higher likelihood of functional significance of the variant.

